# Study protocol and statistical analysis plan for a randomized, double-blind, controlled trial evaluating recombinant human platelet-derived growth factor B (rhPDGF) in the reconstruction of complex head or neck defects following skin cancer excision

**DOI:** 10.64898/2026.05.01.26352276

**Authors:** Cyndi R Clark, Bryan S Blette, Ricardo A Torres Guzman, Melissa D Lempicki, Georgios Karamitros, Franklin Gergoudis, Barite W Gutama, Daniel R O’Neill, Benjamin Savitz, Jacob Smith, Jana K Shirey-Rice, Jill M Pulley, Samuel E Lynch, Trey W McGonigle, Wesley P Thayer

## Abstract

**Background:** Phenome-wide association studies (PheWAS) can reveal novel associations between variants in drug-target genes and disease and, as such, can be used to predict new drug-indication pairs for repurposing drugs with a known mechanism of action. A platelet-derived growth factor receptor beta (PDGFRβ) PheWAS demonstrated that patients with a single nucleotide variant that reduces PDGFRβ expression exhibit a higher prevalence of chronic skin ulcers, skin grafts, and reconstructive surgeries. Recombinant human platelet derived growth factor BB (rhPDGF) is a therapeutic that binds to and activates PDGFRβ and has received FDA approval for multiple indications, including improving healing of lower extremity diabetic neuropathic ulcers, augmenting periodontal bone and soft tissue reconstruction, and stimulating orthopedic bone regeneration. Leveraging a drug-repurposing methodology informed by PheWAS, we hypothesize that rhPDGF will provide therapeutic benefit in the treatment of other complex wounds, like full-thickness surgical wounds of the head or neck that cannot heal by primary intention following skin cancer excision.

**Methods:** This prospective, double-blinded, single-site study aims to enroll 40 participants, randomized at a ratio of 1:1, comparing the efficacy of an advanced wound matrix saturated with rhPDGF or saline. Comparisons will be stratified by anatomical location (scalp/forehead versus face/neck) and maximum surgical defect dimensions (≤ 3cm versus > 3cm). The primary outcome of this study will evaluate the time in days to 81-100% granulation of the wound bed by expert clinical assessment of daily photographs. Secondary outcomes will assess the superiority of the rhPDGF-enhanced wound matrix relative to control with respect to wound granulation rate, epithelialization, complete wound healing, and patient reported outcomes (PROMs).

**Discussion:** Although reconstructive techniques are available for healing complex head and neck wounds following skin cancer excision, these procedures are invasive, and older, frail patients are often suboptimal candidates. There remains a need for less invasive therapeutic approaches that reduce the healing time and mitigate the morbidity associated with chronic wounds. A PheWAS analysis identified complex wounds requiring reconstructive surgery as a novel drug-indication pair for repurposing rhPDGF. This protocol is designed to evaluate the efficacy of an rhPDGF-enhanced advanced wound matrix for healing complex head and neck wounds post skin cancer excision that cannot heal by primary intention.

**Clinical trial registration:** This trial is registered at ClinicalTrials.gov (NCT06634030).

## Introduction

Phenome-wide association studies (PheWAS) are used to identify shared biological mechanisms across multiple diseases and conditions allowing for the identification of new drug indications and targets. These studies leverage electronic health record (EHR)-linked genetic data to identify associations between specific gene variants and aggregated diagnostic billing codes, or PheCodes. If FDA-approved drugs targeting a specific variant gene, protein, or pathway exist, PheWAS can predict novel drug-indication pairs, beyond the drug’s originally intended use. This is achieved by identifying phenotypes, derived from the PheCodes, that occur with a higher or lower prevalence among variant carriers compared to controls. These already approved drugs benefit from established safety data, are frequently available in cost-effective generic formulations, and offer significant advantages for accelerating development timelines through drug repurposing pathways.

Vanderbilt Health’s (VH) academic drug repurposing program leverages PheWAS to identify and validate associations between single nucleotide variants (SNVs) that result in specific protein alterations and disease phenotypes that may be amendable to targeted therapy (1–3). A PheWAS analysis of the platelet-derived growth receptor beta gene (*PDGFRB)* identified an association between SNV rs41287112, which decreases PDGFRβ expression, and an *increased* prevalence of chronic skin ulcers, skin grafts, and reconstructive surgeries in variant carriers. These findings suggest that targeting PDGFRβ may benefit treatment of complex wounds (4). Recombinant human PDGF-BB (rhPDGF) is a pharmaceutical therapeutic that binds and activates PDGFRβ, promoting chemotaxis, mitogenesis, and extracellular matrix deposition (5–7). It is FDA-approved for the treatment of lower extremity diabetic ulcers, periodontal bone and soft tissue reconstruction, and orthopedic bone regeneration with over 150 published clinical studies demonstrating its strong safety profile (8). Leveraging a PheWAS-driven drug repurposing strategy, we hypothesize rhPDGF therapy may be beneficial for non-diabetic, poorly-healing, complex wounds.

Skin cancer, including Basal Cell Carcinoma (BCC), Squamous Cell Carcinoma (SCC), and melanoma, is the most common cancer in the United States, affecting approximately one in five Americans during their lifetime (9–11) . Skin cancer lesions are commonly found on the head and neck due to chronic ultraviolet (UV) radiation exposure and are commonly treated by Mohs micrograph surgery (for BCC and SCC) or wide local excision (for melanoma). These surgical approaches aim to achieve complete tumor excision while preserving healthy tissue and minimizing cosmetic deformities (12,13). In certain cases, full-thickness excision is required due to deep tumor invasion, and postoperative complications can be of concern depending on the tumor size, location, and depth, and the involvement of underlying structures such as muscle, fat, bone, or cartilage (14). Wound healing after surgery is particularly challenging when the margins cannot be approximated with staples, stitches, or glues and the defect is left to heal naturally by secondary intention. In retrospective studies of patients with complex surgical defects of the scalp and forehead with exposed bone, secondary intention healing took 13 to 26 weeks to fully re-epithelialize (15,16), increasing the chances of complications such as infection, necrosis, and unsightly scarring which are commonly associated with these complex wounds.

Rotational flaps and free flaps are reconstructive techniques where larger amounts of tissue with an intact blood supply are taken from a donor site on the patient and used to reconstruct a complex defect. However, older, frail patients are particularly vulnerable to complications and morbidity resulting from these invasive techniques and are often poor candidates for these surgical interventions. Recently, allografts, autographs, and xenografts made of extracellular matrix, cultured skin cells, or synthetic nanofibers have been reported as beneficial for the postoperative management of complex defects by providing protection against infection, promoting granulation and healing (17–21). These monotherapies that mimic individual components of the normal wound healing microenvironment show promise, but there is room for improvement with adjunct therapies that stimulate the signaling factors necessary for wound healing.

A PheWAS identified poorly-healing, complex wounds requiring reconstructive surgery as a novel drug-indication pair for repurposing rhPDGF. Reconstructive surgery for full-thickness surgical defects of the head and neck after skin cancer excision, which often result in exposed bone, is invasive and often challenging in the older population, resulting in increased risk of infection and necrosis. Non-invasive therapies that stimulate wound healing are an unmet need. We propose that rhPDGF will benefit healing of complex head and neck wounds by stimulating chemotaxis and mitogenesis of the cell types that deposit connective tissue, form new blood vessels, and regrow damaged tissue. This protocol will evaluate a rhPDGF-enhanced advanced wound matrix for complete healing of complex head and neck wounds that cannot heal by primary intention and have been referred for reconstructive surgery post skin cancer removal.

## Methods

### Trial design

This study will be carried out in accordance with International Conference on Harmonisation Good Clinical Practice (ICH GCP) and the United States (US) Code of Federal Regulations (CFR) applicable to clinical studies (45 CFR Part 46, 21 CFR Part 50, 21 CFR Part 56, 21 CFR Part 312, and/or 21 CFR Part 812). This Phase II clinical trial will compare the efficacy of an advanced wound matrix soaked in rhPDGF versus saline (control) to heal complex wounds of the head and neck whose margins cannot be approximated to heal by primary intention following skin cancer excision. This prospective, double-blinded, single-site study will randomize participants into two arms – intervention and control – comparing wound bed granulation, epithelialization, complete healing, and scarring and patient reported outcomes (PROMs), including pain and quality of life. The comparisons will also be stratified by anatomical location (scalp/forehead versus face/neck) and greatest dimension (≤ 3cm versus > 3cm) of the surgical defect. To achieve balance in treatment allocation, randomization blocks of 4 (2 interventions: 2 controls) for each stratification group will be employed.

After recruiting, consenting, and screening, eligible participants will undergo the baseline procedure to place the rhPDGF- or saline-saturated wound matrix into the wound bed. Following the baseline procedure, participants will return for their first follow-up visit on day 7 for removal of the foam bolster, clinical examination, photographs, and placement of standard wound dressing. Pain and adverse events will be evaluated and documented weekly through the participants’ electronic health records, phone calls, email surveys, or in-person visits. Participants will return for in-person study visits at weeks 4 and 8 for a clinical exam, photographs, and outcomes assessments and submit daily photographs of the wound while performing dressing changes at home starting on day 7 until day 56. The photographs will be taken using the NetHealth Tissue Analytics imaging application and automatically uploaded to the online platform. Blinded wound experts will analyze the photos, retrospectively, to determine the precise day that the wound bed achieved 81-100% granulation. The NetHealth Tissue Analytics imaging software will also be used to trace and measure granulation, epithelialization, and wound closure rates. Under routine care with a wound matrix (no rhPDGF), the average time to 81-100% granulation is 4-6 weeks in this patient population. Here, we aim to reduce the time to 81-100% granulation by adding rhPDGF.

The Standard Protocol Items: Recommendations for Interventional Trials (SPIRIT) schedule of enrollment, intervention, and assessments is included as **Figure 1**. A graphic study timeline of events is provided in **Figure 2**. The SPIRIT checklist is included in **S1 Appendix**. A full accounting of evaluations and unabridged protocol approved by the IRB is available in **S2 Appendix** (December 19, 2025; version 7.0) and important protocol modifications will be available from the corresponding author and by viewing the ClinicalTrials.gov (NCT06634030) study entry. A written sample consent form approved by the VH IRB is included in **S3 Appendix**.

**Figure 1.**
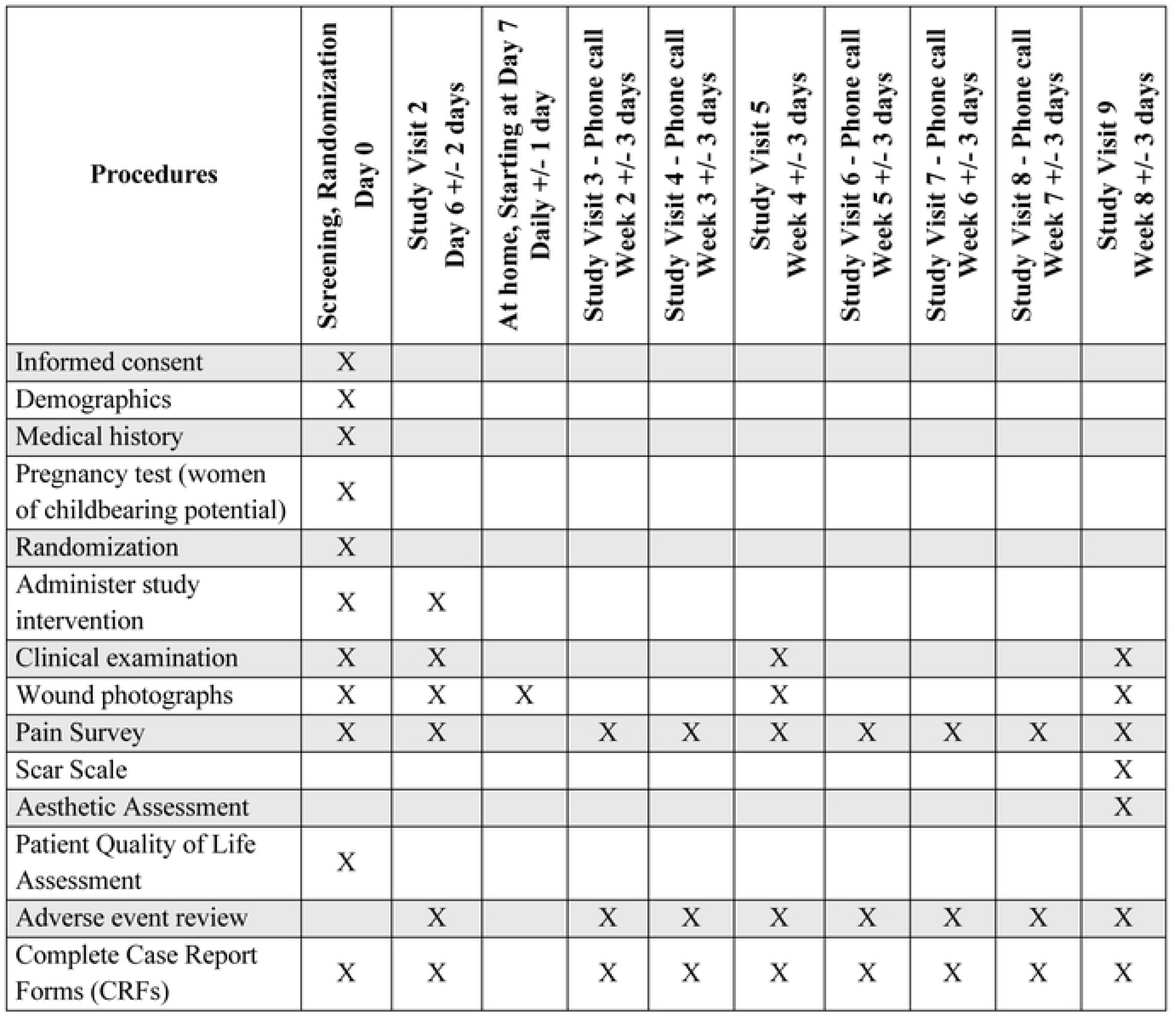
SPIRIT schedule of enrollemnt, intervention, and assessments

**Figure 2.**
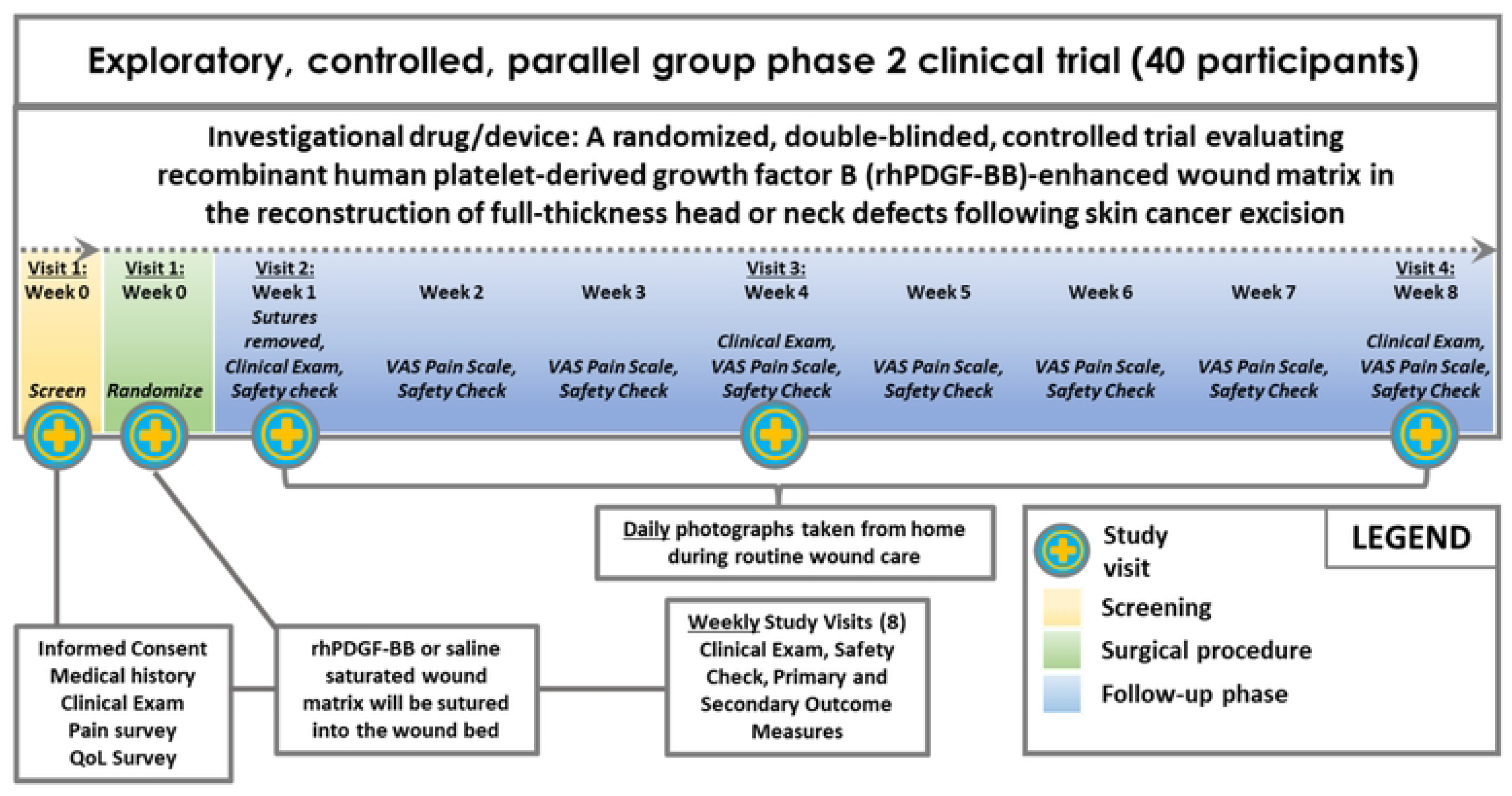
Schematic of study design

### Setting

The trial will be conducted at Vanderbilt Health (VH) in Nashville, TN in the Department of Plastic Surgery.

### Recruitment and consent

This study aims to enroll 40 participants, 20 per arm, that are adults >21 years of age regardless of sex, race, or ethnicity. There is no information currently available regarding differential agent effects in participants defined by race or ethnicity. No vulnerable populations are expected to be enrolled in this study. The target number of patients to be recruited for enrollment is 200 with the expectation of consenting and randomizing ∼25% of those and accounting for discontinuation, withdrawal, and loss to follow-up. This will be a single-site study at VH with an anticipated accrual rate of 1-2 participants per week.

Patients who have recently undergone Mohs surgery or wide local excision to remove skin cancer from the head or neck are referred to the PI for reconstructive surgery when the procedure leaves a complex surgical defect that cannot heal by primary intention. At the initial visit, patients that are determined to be eligible and interested in the trial will sign informed consent and undergo the baseline examination. Informed consent is initiated prior to the individual’s agreeing to participate in the study and continues throughout the individual’s study participation. Informed Consent Forms (ICF) are VH Institutional Review Board (IRB)-approved. Participants who signed consent but did not receive treatment will not be followed further.

### Participants

#### Inclusion criteria

- Underwent surgery to remove skin cancer, either Mohs micrographic or wide local excision, that left a full-thickness surgical defect of the head or neck measuring between 1.5-10 cm in the greatest dimension with negative margins as determined by the Mohs surgeon or the pathology report following excision.
- Margins of the wound cannot be approximated or closed with stitches, sutures, staples, or glue
- Surgeon does not plan for immediate skin graft or flap
- Aged >21 years old
- Willing and able to provide informed consent and comply with the study protocol
- Stated willingness to comply with all study procedures and availability for the duration of the study

#### Exclusion criteria

- Clinically significant medical conditions that would, in the opinion of the investigator, compromise the safety of the participant with study participation and/or the ability of the participant to follow study protocol
- The device will not fit the contour of the base of the wound bed
- Has evidence of current clinical infection as demonstrated by the invasion of bacteria into the healthy viable tissue on the periphery of the wound (colonization of wound bed due to normal flora or environment is not exclusionary)
- Prior radiation therapy at the application site
- Known allergic reactions to porcine tissue, porcine collagen, or yeast-derived products
- Currently enrolled in a drug or device trial or within 30 days of last investigational drug or device administration at baseline where investigational treatment (drug or device) was placed in wound bed or may potentially interact with study treatment
- Women who are known to be pregnant, breastfeeding, or planning to become pregnant during the trial

#### Screen failures

Screen failures are defined as participants who consent to participate in the clinical trial but are not subsequently randomized or for whom the wound matrix is unable to be placed in the wound bed. A minimal set of screen failure information is required to ensure transparent reporting of screen failure participants, to meet the Consolidated Standards of Reporting Trials (CONSORT) publishing requirements and to respond to queries from regulatory authorities. Minimal information includes demography, screen failure details, eligibility criteria, and any serious adverse events (SAE).

Individuals who consent and do not meet the criteria for participation in this trial (screen failure) because of the following criteria may be rescreened once the below criteria have been resolved.

- Pathology report indicates margins aren’t yet clear
- Have an active infection
- Participation in an investigational study as outlined in exclusion criteria
- Pregnant or breastfeeding

### Randomization and blinding

Randomization will occur at the baseline visit at a ratio of 1:1 intervention to control. To achieve balance in treatment allocation, randomization blocks of 4 (1:1 intervention to control) will be stratified by anatomical location, scalp/forehead versus face/neck, and greatest dimension, </= 3cm versus > 3cm, of the surgical defect. Minor imbalance in treatment arm size may occur if there are incomplete randomization blocks. Blinding of the patients, investigators, and study team will be employed to reduce bias in conducting study activities and evaluations. The only personnel that will be unblinded will be an unblinded statistician and the scrub nurse that prepares the wound matrix, by soaking it in either rhPDGF or saline in the operating room. Once prepared, the wound matrix soaked in rhPDGF is indistinguishable from the wound matrix soaked in saline. In the case of a serious adverse event or medical emergency in which knowledge of the treatment is necessary for the participant’s well-being or medical care, the investigator will be unblinded to ensure appropriate medical management.

### Intervention

rhPDGF is highly purified, aseptically processed, and sterile filled into transparent glass syringes. One syringe contains a 0.5 mL solution of 0.3 mg/mL rhPDGF in 20 mM sodium acetate buffer pH6+/-0.5. The wound matrix is available in multiple sizes and can be trimmed or sutured together as needed. For one 4x2cm wound matrix, 4 mL, or 8 syringes, of rhPDGF (1.2 mg) will be used to saturate the matrix in a sterile dish, and multiple wound matrices may be used depending on the size of the wound. All components of the device are biocompatible and absorbable and will only be applied once.

### Administration

In this study the following process will be performed for administration of the intervention:

1. A clinical examination will be conducted to rule out infection, take photographs, and measure the size of the defect.
2. The wound bed will be cleaned and debrided, as needed.
3. The appropriate size wound matrix will be selected based on area of the wound.
4. The matrix will be saturated with the appropriate volume of rhPDGF solution or saline in a sterile dish for at least 10 min.
5. After the matrix is fully saturated, it will be placed into the wound bed and sutured at the margins.
6. The matrix will be trimmed, as needed, to snug fit into the wound bed.
7. The wound bed will be covered with foam bolster that does not get removed during the first week.

### Outcomes

#### Primary outcome

The primary outcome of this study will be to evaluate the potential efficacy of the rhPDGF-enhanced wound matrix on time (in days) to achieve 81-100% granulation of complex head or neck wounds following skin cancer excision. This will be assessed by expert clinical review and assignment of the percent granulation of the wounds in photographs taken daily starting at day 7.

#### Secondary outcomes

The secondary outcomes of this study will assess the effect of the rhPDGF-enhanced wound matrix on wound granulation, epithelialization, complete healing, and aesthetics and participant pain and quality of life after skin cancer excision. Wound granulation rate will be evaluated by percent granulation versus time and weekly ordinal scale outcomes of wound bed granulation (0-20% granulation = 1, 21-40% granulation = 2, 41-60% granulation = 3, 61-80% granulation = 4, 81-100% = 5) as determined by expert clinical review of photographs taken daily starting at day 7. Wound epithelialization will be evaluated by time (in days) to 100% epithelialization and percent epithelialization versus time, and complete healing will be evaluated by time (in days) to complete healing and wound size change over time. Epithelialization and complete healing will be measured utilizing the NetHealth Tissue Analytics software to trace and plot the change in the granulation tissue, new epithelial tissue, and the wound perimeter over time using the daily photographs of the wounds. An aesthetics survey and scar scale will be completed by expert clinicians based on visual assessment, final photograph, and final video completed during the Week 8 follow up visit. PROMs will evaluate change in pain and quality of life over the course of the 8 weeks on study.

#### Feasibility objectives

The feasibility objectives for this study will evaluate the feasibility of recruiting and retaining participants undergoing head and neck reconstruction after skin cancer excision. This will be measured by the total number of participants recruited and eligible, patients randomized per month, percent retention at end of study (8 weeks), and adherence to daily wound photograph schedule.

#### Safety objectives

The safety objectives for this study will evaluate rhPDGF treatment of complex head and neck wounds in patients who underwent skin cancer excision. This will be assessed by qualitative and descriptive summary of all adverse events associated with the intervention, such as but not limited to, wound size assessment, red skin rash around wound area, burning at the application site, swelling at the application site, bleeding at the application site, pain at application site, skin fibrosis around application site, and local recurrence rate of skin cancer.

### Study assessments and procedures

#### Safety assessments

- Clinical examination

`o` At the baseline visit and all follow-up study visits, clinical examination of the defect will visually assess for worsening of wounds including but not limited to infection, excess drainage, chronic inflammation, abnormal bleeding, or fibrosis.
- Patient medical history

`o` At the baseline visit, patients meeting inclusion criteria will be screened for exclusion criteria through their medical history. Participants will review any known allergies, previous surgical procedures, previous complications, and frequency of BCC/SCC occurrence or recurrence.
- Pregnancy test

`o` At the baseline visit, women of childbearing potential will be required to take a pregnancy test and must agree to use two reliable contraceptive methods for the duration of their participation in the study, one of which must be a barrier method
- Assessment of adverse events

`o` The following adverse events will be recorded in the eCRFs and followed until resolution:

∎ Adverse events that are classified as related to study procedures (that is, potentially-, probably-, or definitely related to study procedures or of uncertain relationship to study procedures), regardless of severity
∎ Serious adverse events regardless of relatedness to study procedures.
∎ Clinical adverse events, events diagnosed as clinical conditions, with a severity grade ≥ 3, regardless of relatedness to study procedures.

#### Efficacy assessments

- Clinical examination

`o` At the baseline visit and all follow-up study visits, clinical examination and photographs of the wound bed will be performed to assess the defect size, percent granulation, epithelialization, or wound closure.
- VAS Pain Scale

`o` At the baseline visit and all follow-up study visits, both in-person and by phone, participants will complete the VAS pain scale
- Quality of Life Survey

`o` At the baseline visit and the final study visit (8 weeks), participants will complete the quality of life (QoL) survey.
- Daily Photographs

`o` Starting at Day 7 until the final study visit (8 weeks), participants will take photographs of the wound bed during the daily dressing changes at home.
- Aesthetics Survey

`o` At the 8-week final study visit, the investigator will complete the aesthetics survey.
- Scar Scale

`o` At the 8-week final study visit, the investigator will complete the scar scale.
- Wound Epithelialization

`o` Starting at Day 7 until the wound has closed, the NetHealth Tissue Analytics software will be used to assess the percent wound epithelialization by tracing the epithelial tissue within the wound and tracing the total size of the wound.
- Wound Closure

`o` Starting at Day 7 until the wound has closed, the NetHealth Tissue Analytics software will be used to assess the percent wound closure by tracing the wound in each photograph over time.

### Participant discontinuation/withdrawal from the study

Participants are free to withdraw from participation in the study at any time upon request. An investigator may discontinue or withdraw a participant from the study for the following reasons:

- Failure to heal
- Infection or abscess which requires discontinuation of the study intervention by removal of the wound matrix followed by routine care
- Unacceptable adverse events
- In the judgement of the investigator, further treatment would not be in best interest of patient
- Substantial non-compliance by the patient with the requirements of the study
- The patient uses illicit drugs that may have reasonable chance of interfering with results
- Patient is lost to follow-up
- Development of intercurrent illness or situation which would affect assessments of clinical status and study endpoints to a significant degree
- Pregnancy

`o` In the event a study participant becomes pregnant while on study, the pregnancy will be followed to delivery and maternal and fetal outcomes will be documented.
- Incarceration
- Disease progression which requires discontinuation of the study intervention
- Participant meets an exclusion criterion (either newly developed or not previously recognized) that precludes further study participation

### Risks

The inclusion and exclusion criteria for this study were designed to minimize the risks to participants. The plethora of human safety data that is published supports the expectation that the only risk involved with the use of rhPDGF is the potential to increase acute inflammation at the site of application following intervention. The wound matrix is a biocompatible xenograft that is indicated for the management of wounds. It is contraindicated in individuals that have a known allergy to porcine tissue or active infection at the surgical site. The wound matrix should be used with caution in patients with impaired wound healing due to metabolic disorders, autoimmune diseases, or drug treatment. Potential adverse events that may be observed are those that may occur with any surgery including swelling, bleeding, hematoma, pain, and local inflammation or allergic reaction. The only other risks are associated with routine care procedures. Throughout the life cycle of the study, safety assessments will be used to ensure proper monitoring of adverse events. Safety assessments will consist of clinical examinations, evaluation of patients’ medical history, pregnancy testing, and weekly participant surveys. The potential benefits of this investigational drug and device to improve the healing rates and quality of life far outweigh the minimal risks in individuals with complex defects following skin cancer excision.

### Statistical analysis

Categorical and binary data will be summarized using counts and percentages. Continuous data will be summarized using medians and interquartile ranges. All statistical tests will use an α = 0.05 Type I error rate, using two-tailed p-values as appropriate for the specified hypotheses, but 95% confidence intervals will be given primary focus above p-values. Statistical models will be adjusted for relevant baseline covariates which are predictive of the models’ respective outcomes to improve statistical power. Model assumptions will be checked using visualizations and tests as appropriate, with data transformations and alternative models considered when assumptions are dubious.

#### Statistical hypotheses

- Primary Efficacy Endpoint:

`o` The null hypothesis is that patients in the rhPDGF-enhanced wound matrix arm will have the same time to 81-100% granulation distribution for skin cancer excision surgical defects as patients in the wound matrix saturated with normal saline arm.
`o` The alternative hypothesis is that patients in the rhPDGF-enhanced wound matrix arm will have a different time to 81-100% granulation distribution for skin cancer surgical defects compared to patients in the wound matrix saturated with normal saline arm.
- Secondary Efficacy Endpoints:

`o` The null hypotheses are that rhPDGF-enhanced wound matrix will demonstrate the same average granulation rate, ordinal scale outcomes, time to epithelialization, epithelialization rate, time to complete healing, wound closure rate, pain scale change over time, or patients’ QoL at 8 weeks adjusted for baseline, as the wound matrix saturated with normal saline.
`o` The alternative hypotheses are that rhPDGF-enhanced wound matrix will demonstrate a different average granulation rate, ordinal scale outcomes, time to epithelialization, epithelialization rate, time to complete healing, wound closure rate, pain scale change over time, or patients’ QoL at 8 weeks adjusted for baseline, as the wound matrix saturated with normal saline.

#### Sample size

This study will aim to randomize 40 participants in a 1:1 ratio (20 per arm). To achieve balance in treatment allocation, randomization blocks of 4 (2 interventions : 2 controls) will be stratified by anatomical location, scalp/forehead versus face/neck, and greatest dimension, ≤ 3cm versus > 3cm, of the surgical defect. Time to readiness for a skin graft was used for power calculations; this outcome was replaced in a revision of the study protocol with time to 81-100% granulation of the wound, a more appropriate measure of healing that still aligns with our power calculation assumptions. Based on the prior work in the literature, we assume that patients in the placebo arm will have a median time to 81-100% granulation of the wound of 28 days. Assuming that both arms follow an exponential distribution for this primary outcome and that the intervention arm will have a median time to 81-100% granulation of 14 days, a sample size of 20 participants per arm achieves greater than 50% power to reject a log-rank test (22) corresponding to hypotheses outlined above. We assumed that there would be 5% dropout before end of study in each arm, with uniform dropout over time. Power calculations were performed in R version 4.3.1 (23). Primary analyses will adjust for baseline covariates, which may further improve statistical power. Power across a range of assumptions was calculated and displayed in **Table 1**.

**Table 1.** Power for detection of differences in the primary outcome (time to readiness for skin grafting or 81-100% granulation) between the intervention group and placebo group, assuming 20 participants enrolled in both groups.

| Observed median time to readiness for skin graft or 81-100% granulation (days) |  | Power to detect difference |
| --- | --- | --- |
| Intervention Group | Placebo Group |  |
| 7 days | 28 days | 98% |
| 10 days | 28 days | 84% |
| 14 days | 28 days | 52% |
| 21 days | 28 days | 13% |
| 7 days | 35 days | >99% |
| 10 days | 35 days | 94% |
| 14 days | 35 days | 74% |
| 21 days | 35 days | 29% |
| 28 days | 35 days | 10% |

#### Primary outcome analysis

The primary endpoint is time to 81-100% granulation of the wound based on expert assessment of patient wound photos. The time will be in days based on when both expert reviewers agree on the granulation reaching 81-100% granulation by independent review. This time-to-event outcome will be modeled using a Cox proportional hazards model, conditioning on the treatment arm and relevant baseline covariates, as outlined in the SAP. Patients who drop out before achieving 81-100% granulation of the wound will be treated as censored, which will be assumed non-informative. The effect will be presented as an adjusted hazards ratio with a 95% confidence interval, as well as the marginal hazard ratio after performing standardization. A p-value less than 0.05 will be considered statistically significant, but interpretation will be primarily focused on the 95% confidence interval. Kaplan-Meier plots will be used to visualize the results and assess the proportional hazards assumption. Missing data are expected to be limited but will be handled using multiple imputation if there are insufficient photos to determine a precise day where the primary outcome is achieved. Sensitivity analysis will include individuals who are lost to follow-up before achieving 81-100% granulation of the wound, considering multiple imputations for their 81-100% granulation time based on their trajectory of healing before dropout. The primary analysis will be intention-to-treat (ITT) based on randomized arm, as the investigational product is not available to patients who are not randomized to treatment. Supplementary Bayesian analyses will be used to quantify probability of patient benefit.

#### Secondary outcomes analysis

The secondary efficacy endpoints will be analyzed for all randomized participants. Analyses will not be dependent on the findings of the primary endpoint.

The first secondary endpoint is percent granulation over time as measured from daily photos. This endpoint will be rated by two adjudicators and averaged. This endpoint will be analyzed using a longitudinal cumulative probability ordinal regression model, which has been shown to perform well with continuous outcomes (24) and can handle the truncated nature of the outcome variable. The model will condition on the treatment arm and relevant baseline covariates, including a random intercept at the individual level to account for within-individual correlation. Time trends will be modeled using spline functions. The treatment effect results will be presented as an adjusted odds ratio and a marginal odds ratio after performing standardization. The proportional odds assumption will be made and assessed by comparing fitted and observed probabilities under proportional and partial proportional odds; the assumption will be relaxed if the assessment indicates lack of fit. Missing data are handled naturally by mixed-effects models fit by maximum likelihood under a missing at random assumption. If missingness is substantially more than expected, alternative methods such as multiple imputation will be considered. The next secondary endpoint of weekly ordinal scale outcomes will be modeled using the same framework and approach.

The next secondary endpoints are time to epithelialization and time to complete healing based expert review and tracing of epithelial tissue and total wound size using the Tissue Analytics software. These will both be analyzed using the same approach as the primary outcome.

The next endpoint is percent epithelialization over time and wound closure over time measured from the tracings in the photos using the Tissue Analytics software. This will be analyzed using a cumulative probability ordinal regression model, as described for the similar longitudinal percent granulation variables. Longitudinal pain scale will also be modeled in the same framework.

These models will be further adjusted for corresponding baseline outcome values. Missing data will be handled by the mixed-effects models as described for the percent granulation endpoint.

Ordinal participant quality of life responses, aesthetics survey variables, and scar scale based on final photos at 8 weeks will be analyzed in the same framework without the random intercept, as outcomes are only collected at baseline and 8 weeks. Models will be adjusted for corresponding baseline outcome variables. Missing outcomes at 8 weeks will not be imputed for those lost to follow-up.

The stratification variables for randomization will be adjusted for as baseline covariates in all models unless the number of degrees of freedom available in a given model prohibits this. Subgroup analyses in the strata defined by these variables will be conducted for wound healing outcomes.

#### Safety analyses

Monitoring and reporting of safety events will be conducted continuously as described in the Data and Safety Monitoring Plan. This section describes the assessment of safety endpoints at the time of final analysis. The frequencies of study-related adverse events and all other safety endpoints will be reported along with the treatment effect on the odds of these events (i.e., the odds ratio) and associated 95% confidence intervals using logistic regression for binary safety outcomes and the same methods for ordinal safety outcomes as those outlined for ordinal secondary endpoints above. Descriptive summaries of all adverse events will be provided.

### Ethics

Participant confidentiality and privacy is strictly held in trust by the study team, and all research activities will be conducted in as private a setting as possible. This confidentiality is extended to cover the participant’s clinical information and the study protocol, documentation, data, and all other information generated. No information concerning the study or data will be released to any unauthorized third party. All faculty and staff working on the study receive and provide documentation of training in Good Clinical Practice (GCP) as part of their onboarding and continuing education.

Data, safety, and enrollment monitoring is conducted to ensure that the rights and well-being of trial participants are protected, that the reported trial data are accurate, complete, and verifiable, and that the conduct of the trial is in compliance with the currently approved protocol/amendment(s), with International Conference on Harmonisation Good Clinical Practice (ICH GCP), and with applicable regulatory requirement(s). Safety oversight will be under the direction of a Data and Safety Monitoring Board (DSMB) composed of 3 independent members. They will be experts/representatives such as clinicians, clinical trialists, and statisticians. They will not be study investigators or involved in execution of the study protocol. Furthermore, no member should have financial, proprietary, professional, or other interests that may affect impartial, independent decision-making by the DSMB.

This study will comply with the NIH Data Sharing Policy and Policy on the Dissemination of NIH-Funded Clinical Trial Information and the Clinical Trials Registration and Results Information Submission rule. As such, this trial will be registered at ClinicalTrials.gov, and results from this trial will be submitted to ClinicalTrials.gov. In addition, every attempt will be made to publish results in peer-reviewed journals.

### Trial status

Screening, enrollment, and randomization for this trial began in May 2025. Participant screening and enrollment will end June 2026. Data collection will be completed in August 2026, and results will be reported shortly after. The trial is still ongoing at the time of submission.

## Discussion

Using a PheWAS-leveraged methodology, we identified a novel drug-indication pair for a repurposed FDA-approved therapy with a strong safety profile and extensive history of human use. Through Vanderbilt Health’s de-identified electronic health records database, the Synthetic Derivative (SD), in conjunction with a biobank of deidentified patient DNA samples with SNV genotyping data, BioVU (1–3), PheWAS revealed that patients carrying SNV rs41287112, which decreases *PDGFRB* expression, exhibit a significantly higher prevalence of chronic ulcers, skin grafts, and reconstructive surgeries. PDGFRβ is a tyrosine kinase receptor that binds endogenous platelet-derived growth factors to activate and regulate the healing cascade, but the levels of these endogenous proteins can vary depending on genetics, age, and overall health. Recombinant human PDGF (rhPDGF) is a therapeutic administered at controlled concentrations to recapitulate endogenous growth factor activity by binding to PDGFRβ, initiating the healing cascade, and promoting a positive feedback loop to increase receptor expression. rhPDGF is currently approved for treatment of lower extremity diabetic ulcers and as an adjunct for periodontal and orthopedic tissue regeneration. We hypothesize that rhPDGF will provide therapeutic benefit in other non-healing, complex wounds that would otherwise require reconstructive surgery.

Healing large complex wounds that cannot be closed with staples or sutures remains challenging, particularly in older patient populations, who have a significantly higher incidence of skin cancers. These patients are suboptimal candidates for invasive reconstructive surgeries, thereby increasing the risks of complications, including infection, necrosis, and unsightly scarring of the wound. Synthetic and animal-sourced skin substitutes have improved healing rates, provide protection, and offer mechanical support for complex wounds by mimicking the structural organization of human skin and providing an optimized environment for cell migration, attachment, and vascularization (25–27); however, further advancements are still needed to reduce the healing time and improve the aesthetics and integrity of the scar. Preclinical and clinical studies have demonstrated synergistic results with mesenchymal precursors, platelet-rich plasma, and growth factor proteins incubated with bioabsorbable substrate with respect to wound healing (28). More recently, dehydrated placental tissue allografts comprised of extracellular matrix (ECM) proteins and growth factors, as well as autologous platelet rich plasma (PRP) containing cytokines, chemokines, and growth factors were evaluated for post-Mohs chronic wound healing, however more studies are required to better define their efficacy an optimal dosing strategies (29–32). Together, our PheWAS findings for *PDGFRB,* in combinations with a review of the existing literature, support the feasibility of designing a clinical trial to evaluate the effectiveness of a rhPDGF-enhanced advanced wound matrix for complete healing of complex head and neck wounds after skin cancer excision.

Several challenges and limitations with this clinical trial must be acknowledged. Underlying tissue composition, such as muscle, fat, bone, and cartilage, as well as wound size, significantly influence the wound healing timeline. To account for these sources of variability, outcome assessments will include stratified subgroup analyses for anatomical location (scalp/forehead versus face/neck) and greatest dimensions (≤ 3cm versus > 3cm) of the surgical defect. Furthermore, this study requires a high degree of participant engagement, as efficacy outcomes will be analyzed using daily wound photographs obtained at home during dressing changes, in addition to weekly participant surveys. To enhance participant compliance, an application, NetHealth Tissue Analytics, is downloaded to the participants’ mobile device, providing daily reminders and automatic uploads. This platform is designed to be user-friendly and to facilitate consistent data capture. Despite these challenges, if successful, this trial would inform and guide the regulatory pathway for a much-needed alternative treatment option aimed at improving healing outcomes and reducing morbidity associated with complex wounds following skin cancer excision. In addition, the study will further support the utility of a PheWAS-leveraged approach for drug development and repurposing.

## Data Availability

No datasets were generated or analysed during the current study. All relevant data from this study will be made available upon study completion.

## Supporting information

S1 Appendix. SPIRIT checklist

S2 Appendix. Study protocol

S3 Appendix. Consent form

